# Cohort Versus Nested Case-Control Designs for Predicting Early-Onset Gastrointestinal Cancer Using Electronic Health Records

**DOI:** 10.64898/2026.10.16.26365758

**Authors:** Ruochong Fan, Sina Azadnajafabad, Jenna M. Reps, Benjamin Bowe, Mackenzie R. Hofford, Kian-Huat Lim, George Hripcsak, Yin Cao, Linying Zhang

**Author notes:** **Corresponding author**: Linying Zhang, PhD, Institute for Informatics, Data Science and Biostatistics, Washington University School of Medicine in St. Louis, 660 S. Euclid Ave., St. Louis, MO 63110, USA.

## Abstract

**Purpose:** To determine how cohort and case-control study designs affect the discrimination, calibration, and feature rankings of EHR-based models for predicting early-onset gastrointestinal cancer.

**Methods:** Using EHR data from an integrated health system, we developed models to predict EOGIC within 2 years after an outpatient visit among adults age 18-49 years at the time of the visit. We compared two cohort and five case-control designs. Models included L1-regularized logistic regression (LASSO), XGBoost, and multilayer perceptrons (MLP). Models were trained with nested five-fold cross-validation. We assessed model discrimination using area under the precision recall curve (AUPRC), area under the receiver operating characteristic curve (AUROC), calibration before and after isotonic recalibration, and similarity of feature rankings across study designs and predictive models.

**Results:** AUROC does not depend on outcome prevalence, it is suitable for comparing discrimination across study designs with different outcome prevalences; however, it was less sensitive to variation in model performance in this rare-outcome setting. AUPRC estimated in case-enriched samples did not generalize to the incidence-representative population in the intended deployment setting. Raw predicted probabilities substantially overestimated observed risks under both study designs. Isotonic recalibration improved calibration, although residual miscalibration remained. Cohort-based feature rankings were more consistent across algorithms, whereas case-control-based rankings depended more strongly on algorithms.

**Conclusion:** Study design substantially affects the discrimination, calibration, and feature-ranking patterns of EHR-based EOGIC prediction models. Cohort designs are generally better suited to EHR-based models for early detection because they preserve the outcome incidence and temporal relationship between the predictors and the outcome. When case-control designs are used for model development, models should be evaluated and recalibrated in an incidence-representative cohort that reflects the intended clinical setting.

**BACKGROUND AND SIGNIFICANCE:** Early-onset cancers, generally defined as cancers diagnosed before age 50 years, are an increasing public health concern. In the United States, the age standardized incidence of early-onset gastrointestinal cancers (EOGIC), including cancers of the esophagus, stomach, small intestine, cecum, appendix, colon, rectum, liver, intrahepatic bile duct, gallbladder, other biliary organs, and pancreas, increased by approximately 2.2% annually from 2010 to 2019^1^, making it the largest annual increase among early-onset cancer groups.

Despite its rising incidence, EOGIC remains uncommon, with an incidence of up to 15 cases per 100,000 person-years.^1,2^ This low incidence limits the feasibility of population-wide screening. Among all GI cancers, only colorectal cancer screening guidelines recommend screening starting at age 45 years. However, because the median age at EOCRC diagnosis is 44 years^3^, this threshold leaves approximately half of patients below the recommended screening age. Routine screening is not recommended for average-risk individuals for other gastrointestinal cancers. Thus, personalized risk stratification using routinely available clinical information could help identify patients at elevated risk and support early detection.

Electronic health records (EHRs) offer an opportunity to assess EOGIC risk using routinely collected clinical data, including medical history, vital signs, and laboratory measurements.^4,5^ Among early-onset gastrointestinal cancers, EOCRC has been studied most extensively. We identified 13 studies that developed EHR-based risk models for EOCRC between 2011 and 2026, among which 9 studies (69.2%) used case-control designs,^6–14^ and 4 (30.8%) used cohort designs^15–18^, while study-design choices are usually not reported.

Study design can have profound impact on the discriminative performance, risk calibration, and clinical interpretation of predictive models.^19–23^ Previous studies comparing cohort and case-control designs have shown that case-control sampling can lead to miscalibrated absolute risk estimates.^19,23^ Case-control design, by indexing predictors relative to the event date can introduce “temporal bias,” that is, predictors measured close to the event enhance discrimination under retrospective evaluation but may not generalize to prospectively deployable settings.^24^ Evidence to guide the selection of study designs for prediction model development of early-onset cancer remains limited. This gap is important because early-onset cancer is a very rare outcome, creating a strong incentive to enrich cases during model development, yet it is unclear whether performance gains observed under case-control designs persist when models are evaluated prospectively in a population representative of the model deployment setting.

## OBJECTIVE

In this study, we compared two cohort and five NCC designs for EHR-based EOGIC prediction. To assess whether the influence of study designs depended on the modeling choices, we evaluated L1-regularized logistic regression (LASSO), eXtreme Gradient Boosting (XGBoost), and multilayer perceptron (MLP) models and compared their discrimination, calibration, temporal transportability, and feature importance patterns. We also assessed whether recalibration using a deployment-aligned cohort could improve the alignment of predicted and observed risks. Our objective was to inform the selection of study designs and modeling approaches for identifying patients at elevated EOGIC risk using routinely collected EHR data.

## MATERIALS AND METHODS

### Data Source

This was a retrospective study using EHR data, extracted from the Washington University in St. Louis Research Data Core (RDC), a repository of patient clinical data originating from the BJC HealthCare and Washington University Physicians Epic EHR system. The EHR database was standardized to the OMOP Common Data Model version 5.3.^25,26^ This study was approved by the Washington University Institutional Review Board (#202510130) and granted a waiver of HIPAA Authorization for the use of Protected Health Information (PHI), hence participant consent was not required.

### Outcome and Features

The outcome was incident EOGIC, including primary malignant neoplasms of the esophagus, stomach, small intestine, colon, rectum, rectosigmoid junction, anal canal, liver, gallbladder, biliary tract, and pancreas. Patients were required to have at least two qualifying diagnosis codes to be considered as cases. The date of the first qualifying cancer diagnosis was used as the diagnosis date. The code set for defining EOGIC was available in Supplementary Table 5.

Features include age, sex, race, clinical conditions, and laboratory test results, yielding 16,559 features. For all study designs, features were extracted from data recorded during the 365 days prior to the index date. Sex, race, and clinical conditions were represented as binary indicators; age was represented using categorical age-bin indicators. Recorded laboratory measurements were unit-standardized and min-max normalized using parameters estimated from the training data. Missing measurements were coded as zero and a binary missing indicator was included to encode missing laboratory measurements.

### Study Designs and Populations

We compared two cohort designs and five NCC designs (Figure 1). These designs differed in how they defined the index date and the population at risk.

**Figure 1.**
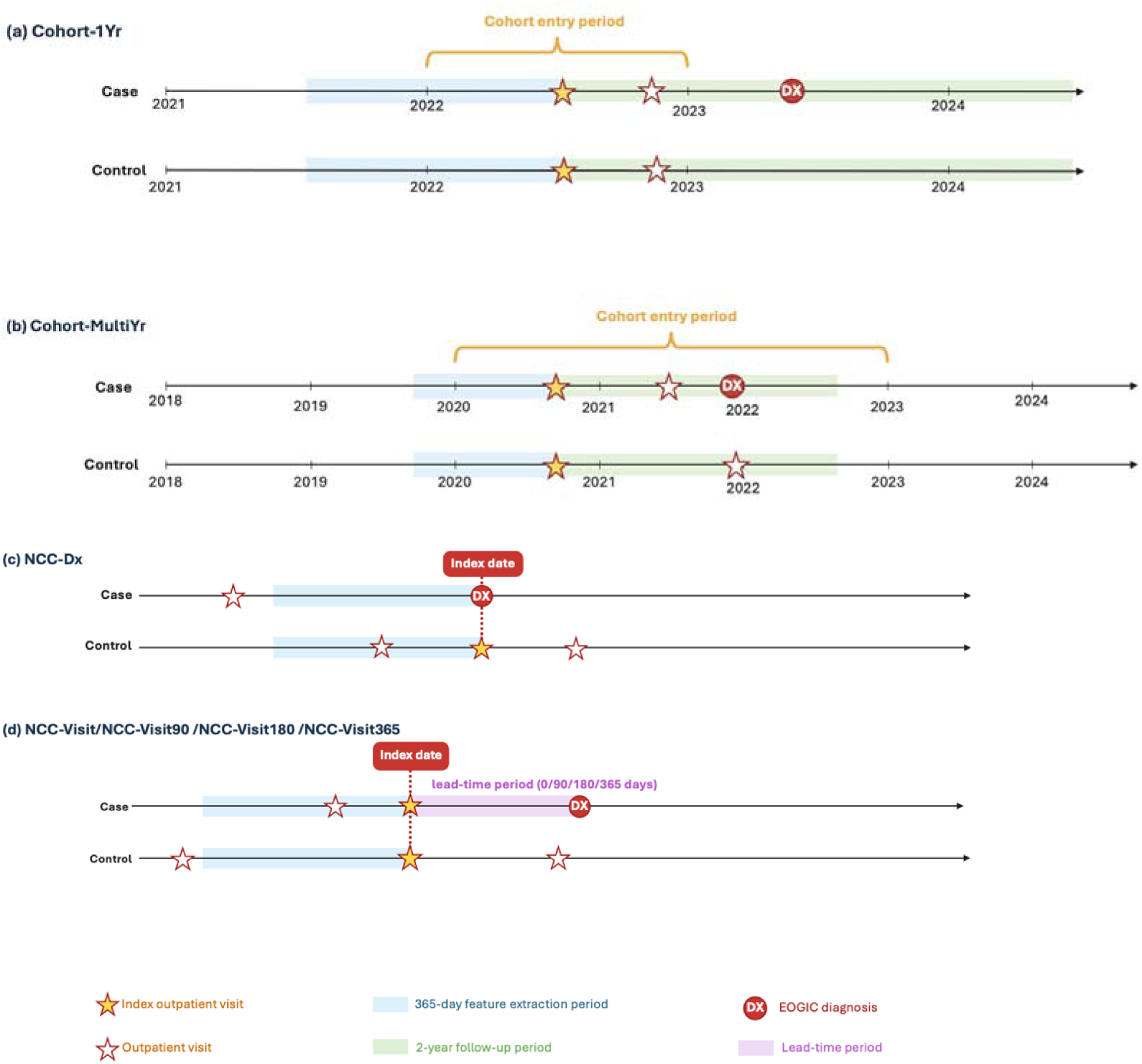
Illustration of the cohort and nested case-control study designs. (A) In the cohort designs, the first qualifying outpatient visit served as the index date, predictors were ascertained during the preceding 365 days, and incident EOGIC was assessed during two years of follow-up. (B) In the diagnosis-anchored nested case-control design (NCC-Dx), the first EOGIC diagnosis served as the case index date. (C) In the visit-anchored designs, the case index date was an outpatient visit occurring at least 0, 90, 180, or 365 days before diagnosis. In the NCC designs, controls were sampled at a 1:10 case-to-control ratio and matched on age, sex, and index calendar date. EOGIC, early-onset gastrointestinal cancer; NCC, nested case-control; NMSC, nonmelanoma skin cancer.

Because eligibility was assessed relative to the index date, which differed across study designs, the study population was constructed separately for each design. Unless otherwise specified, eligible patients were 18-49 years of age at the design-specific index date, had at least 365 days of continuous observation before that date, and had no prior cancer diagnosis other than nonmelanoma skin cancer.

**Cohort designs.** We implemented two cohort variants with different cohort entry periods. In the one-year cohort (cohort-1Yr), eligible patients were those with at least one outpatient visit in 2022, and the first qualifying visit in that year served as the index date. In the multi-year cohort (cohort-MultiYr), eligible patients were those with at least one outpatient visit between January 1, 2020 and December 31, 2022, and the first qualifying visit during this three-year period served as the index date. In both designs, cases were patients who developed EOGIC within two years after the index date, whereas controls remained free of EOGIC throughout the two-year follow-up period.

**Nested case-control designs.** The source cohort comprised all patients with at least one outpatient visit recorded in the EHR database.

We evaluated two index-date anchoring strategies. In the diagnosis-anchored NCC (NCC-Dx) design, the index date for cases was the first EOGIC diagnosis in the patient’s EHR. All cases were required to meet the eligibility criteria described above. The control index dates were assigned as described below.

In the visit-anchored NCC design, the index date for cases was an outpatient visit prior to the EOGIC diagnosis. To examine the impact of potential diagnostic workup signals occurring close to diagnosis, we implemented four visit-anchored NCC designs with different lead-time restrictions between the index visit and the diagnosis. In NCC-visit, NCC-visit90, NCC-visit180, and NCC-visit365, the index date was the outpatient visit with a minimum lead time of 0, 90, 180, and 365 days prior to diagnosis, respectively. At the index date, cases were required to meet all eligibility criteria described above, including at least 365 days of continuous observation. Consequently, the total required observation period before diagnosis increased as the minimum lead time increased.

Across all NCC designs, controls were sampled for each case at a 1:10 case-to-control ratio from members of the source cohort who remained free of EOGIC at the corresponding case index date. Controls were matched on age and sex. They were required to have an outpatient visit preceding the case index date. The qualifying outpatient visit served as the control index date, at which all eligibility criteria were met.

### Temporal Cohort Definition

We constructed a temporal evaluation cohort using the same eligibility and outcome definitions as cohort-1Yr design, with cohort entry period shifted to 2023. Cases were patients who developed EOGIC within two years after the index date, whereas controls remained free of EOGIC throughout the two-year follow-up period. This cohort was used to assess model transportability in future patient population from the intended deployment setting.

### Model Training

We compared LASSO, XGBoost, and MLP, representing linear, tree-based, and neural-network approaches, respectively.

Models were trained separately within each study design using stratified nested five-fold cross-validation. Each outer fold served once for evaluation, while the remaining folds underwent five-fold inner cross-validation for hyperparameter selection based on AUPRC. The selected configuration was refitted on the outer training folds and evaluated on the held-out fold.

For LASSO, the regularization parameter was selected from a prespecified grid of candidate penalty values (1e-5, 3e-5, 1e-4, 3e-4, and 1e-3) within the inner cross-validation loop. Because the outcome was rare, class imbalance was addressed by applying class weights that increased the contribution of case observations relative to controls.

For XGBoost the hyperparameter search included maximum tree depth of 6 or 10, minimum child weight of 1.0 or 10.0, and with or without class weighting. The number of trees was fixed at 500 trees, the subsampling fraction and column subsampling fraction per node were both fixed at 0.8, and the learning rate was fixed at 1.0.

For MLP, we trained fully connected neural networks using the same design-specific tabular feature matrices. We tuned hyperparameters of hidden-layer architectures, dropout rates, and learning rates. Models were optimized using AdamW and weighted binary cross-entropy loss, with the positive-class weight calculated as the ratio of noncases to cases within the corresponding training subset. The MLP hyperparameter search space and design-specific selection patterns are summarized in Supplementary Table 3.

### Model Recalibration

Post hoc isotonic calibration was performed separately by model and study design.^27^ For models trained using cohort designs, we fitted the isotonic recalibration function using data from the corresponding outer validation fold. For models trained using NCC designs, the cohort-1Yr dataset was used for recalibration. Cohort-based datasets were used for recalibration across all study designs because they preserve the underlying outcome prevalence.

### Evaluation of Discriminative Performance

For internal evaluation, discrimination was assessed using the outer validation fold from each study-design-specific development dataset. For MLP, hyperparameters were selected exclusively within the outer training data through 5-fold inner cross-validation, after which the outer validation fold was used for early stopping and performance evaluation. Performance metrics were summarized across the five outer folds, and confidence intervals were calculated from variation across folds. For temporal evaluation, each model was applied to the temporal cohort, and temporal performance was summarized across the five models from the same study design. We evaluated internal and temporal discrimination using AUROC and AUPRC.^28,29^ We used temporal AUPRC as the primary metric for comparing designs because it summarizes the trade-off between identifying future early-onset gastrointestinal cancer and limiting false alerts at the observed two-year cancer incidence in the outpatient setting.

### Evaluation of Model Calibration

Model calibration was evaluated using Brier scores and calibration plots. Each model was applied to the temporal cohort to generate predicted probabilities. Patients were grouped by predicted probability and mean predicted risk was compared with the observed event rate in each decile. Calibration curves were plotted with the mean predicted probability on the x-axis and observed event rate on the y-axis, with the 45-degree diagonal representing perfect calibration. Brier scores were calculated as the mean squared difference between predicted probabilities and observed binary outcomes, with lower scores indicating better overall prediction accuracy.

### Feature Importance

Feature importance was determined separately for each model. For LASSO, predictors were ranked by the absolute value of their regression coefficients. For XGBoost, predictors were ranked using gain importance, calculated as the average improvement in the model’s loss function across splits using each feature.^30^ For MLP models, feature importance was estimated using gradient × input attribution, a gradient-based attribution approach in which the gradient of the model output with respect to each input feature is multiplied by the observed feature value.^31^ Features were ranked by the mean absolute attribution score across sampled patients and outer cross-validation folds.

For clinical visualization, we displayed the top 20 features from Cohort-MultiYr LASSO and NCC-Dx XGBoost, grouped into descriptive clinical categories. LASSO features were selected by the absolute magnitude of their mean signed coefficients across outer folds, with signs retained. XGBoost features were selected using full-model gain, expressed as a percentage of the sum of gain scores across all features.

To assess the similarity between feature rankings across study designs and algorithms, we used rank-biased overlap (RBO)^32^, which quantifies similarity based on the overlap between two ranked lists at progressively increasing depth. RBO ranges from 0 to 1, with larger values indicating greater agreement. We chose RBO over conventional correlation coefficients because it places greater weight on agreement near the top of the rankings and accommodates rankings of unequal length. We set the persistence parameter to 0.99, yielding a relatively gradual decline in weights and an effective evaluation depth of approximately 100 features. Within each cross-validation fold, we calculated RBO for every pair of algorithm-design combinations. For each pair, RBO values were then averaged across the five outer folds, and the resulting mean similarities were visualized as a heatmap.

## RESULTS

### Study design and population

We evaluated two cohort and five NCC designs (Figure 1). The Cohort-1Yr development cohort included 256,854 patients, with 303 EOGIC cases (2-year cumulative incidence of 0.12%). The Cohort-MultiYr development cohort included 415,529 patients and 711 cases (2-year cumulative incidence of 0.17%). The temporal cohort included 258,642 patients and 311 cases (2-year cumulative incidence of 0.12%) (Table 1; Supplementary Table 1). All NCC datasets had a fixed case fraction of approximately 9% due to 1:10 case-to-control matching ratio, which is much higher than the 0.12%-0.17% cumulative incidence observed in the cohort-based datasets.

**Table 1.** Characteristics of the Cohort-MultiYr development cohort and the temporal evaluation cohort. Values are presented as n (%) unless otherwise specified. Clinical conditions were ascertained during the 365-day predictor window preceding the index date. EOGIC, early-onset gastrointestinal cancer; IQR, interquartile range; SD, standard deviation.

| **Cohort size** |  |  |
| **Characteristic** | **Cohort-MultiYr development cohort** | **Temporal evaluation cohort** |
|---|---|---|
| Total patients | 415,529 | 258,642 |
| EOGIC cases | 711 (0.17%) | 311 (0.12%) |
| **Demographics** |  |  |
| Age, mean (SD) | 33.9 (9.0) | 34.2 (9.0) |
| Age, median [IQR] | 34.0 [26.0, 42.0] | 35.0 [27.0, 42.0] |
| *Age group, years* |  |  |
| 18-24 | 82,641 (19.9%) | 49,942 (19.3%) |
| 25-29 | 61,316 (14.8%) | 34,261 (13.2%) |
| 30-34 | 67,026 (16.1%) | 41,510 (16.0%) |
| 35-39 | 69,254 (16.7%) | 43,734 (16.9%) |
| 40-44 | 72,594 (17.5%) | 48,642 (18.8%) |
| 45-49 | 62,698 (15.1%) | 40,553 (15.7%) |
| *Sex* |  |  |
| Female | 265,825 (64.0%) | 171,760 (66.4%) |
| Male | 149,653 (36.0%) | 86,834 (33.6%) |
| Other/unknown | 51 (0.0%) | 48 (0.0%) |
| *Race* |  |  |
| White | 299,289 (72.0%) | 190,778 (73.8%) |
| Black or African American | 106,123 (25.5%) | 60,927 (23.6%) |
| Asian | 10,117 (2.4%) | 6,937 (2.7%) |
| Other/unknown | 0 (0.0%) | 0 (0.0%) |
| *Ethnicity* |  |  |
| Not Hispanic or Latino | 390,254 (93.9%) | 246,727 (95.4%) |
| Hispanic or Latino | 10,095 (2.4%) | 6,814 (2.6%) |
| Other/unknown | 15,180 (3.7%) | 5,101 (2.0%) |
| **Comorbidities** |  |  |
| *Gastrointestinal and hepatobiliary* |  |  |
| Abdominal pain | 14,663 (3.5%) | 12,569 (4.9%) |
| Anorectal disorder | 909 (0.2%) | 989 (0.4%) |
| Benign neoplasm of ascending colon | 147 (0.0%) | 272 (0.1%) |
| Benign neoplasm of cecum | 103 (0.0%) | 161 (0.1%) |
| Chronic gastritis | 923 (0.2%) | 893 (0.3%) |
| Disease of liver | 915 (0.2%) | 913 (0.4%) |
| Disorder of intestine | 3,849 (0.9%) | 3,535 (1.4%) |
| Disorder of pancreas | 548 (0.1%) | 501 (0.2%) |
| Gastroesophageal reflux disease without esophagitis | 14,537 (3.5%) | 13,874 (5.4%) |
| Hemorrhage of rectum and anus | 1,460 (0.4%) | 1,511 (0.6%) |
| Intestinal obstruction | 495 (0.1%) | 436 (0.2%) |
| *Cardiovascular and metabolic* |  |  |
| Chest pain | 14,523 (3.5%) | 11,120 (4.3%) |
| Essential hypertension | 27,027 (6.5%) | 22,749 (8.8%) |
| Hyperglycemia | 1,728 (0.4%) | 1,873 (0.7%) |
| Hyperlipidemia | 9,067 (2.2%) | 8,274 (3.2%) |
| Morbid obesity | 12,392 (3.0%) | 14,962 (5.8%) |
| Obesity | 14,765 (3.6%) | 16,839 (6.5%) |
| *Mental health and substance use* |  |  |
| Anxiety disorder | 24,314 (5.9%) | 21,825 (8.4%) |
| Depressive disorder | 5,343 (1.3%) | 8,999 (3.5%) |
| Generalized anxiety disorder | 6,979 (1.7%) | 11,627 (4.5%) |
| Nicotine dependence | 25,121 (6.0%) | 11,994 (4.6%) |
| *Respiratory and infectious* |  |  |
| Acute pharyngitis | 9,425 (2.3%) | 10,143 (3.9%) |
| Cough | 11,853 (2.9%) | 9,906 (3.8%) |
| Dyspnea | 11,083 (2.7%) | 7,972 (3.1%) |
| Uncomplicated asthma | 11,656 (2.8%) | 7,051 (2.7%) |
| *Pain and musculoskeletal* |  |  |
| Chronic pain | 12,385 (3.0%) | 12,604 (4.9%) |
| Headache | 12,757 (3.1%) | 9,329 (3.6%) |
| Low back pain | 10,814 (2.6%) | 8,670 (3.4%) |
| *Nutritional* |  |  |
| Severe protein-calorie malnutrition (Gomez: less than 60 percent of standard weight) | 760 (0.2%) | 578 (0.2%) |
| Vitamin D deficiency | 9,938 (2.4%) | 13,611 (5.3%) |

The Cohort-MultiYr development cohort and the temporal validation cohort were similar in age (mean, 33.9 versus 34.2 years) and racial composition, although the temporal cohort included more women (66.4% versus 64.0%). Some conditions, or example, abdominal pain (4.9% versus 3.5%), essential hypertension (8.8% versus 6.5%), obesity (6.5% versus 3.6%), anxiety disorder (8.4% versus 5.9%), and vitamin D deficiency (5.3% versus 2.4%) were more prevalent in the temporal cohort (Table 1).

### Model discrimination

AUPRC and AUROC are shown in Figure 2; additional performance metrics are provided in Supplementary Table 2. Case-control designs had substantially higher internal AUPRCs than cohort designs; however, these higher values did not generalize to the temporal validation cohort. Because AUPRC depends on outcome prevalence, internal AUPRCs should not be directly compared across cohort and case-control designs because these study designs have very difference case prevalences. These findings underscore the importance of evaluating model discrimination in a dataset whose outcome prevalence reflects the intended deployment setting. This can be achieved by using a cohort design for both model development and evaluation or, when necessary, by developing the model using a case-control or other case-enriched design and evaluating it in a dataset constructed using a cohort design.

**Figure 2.**
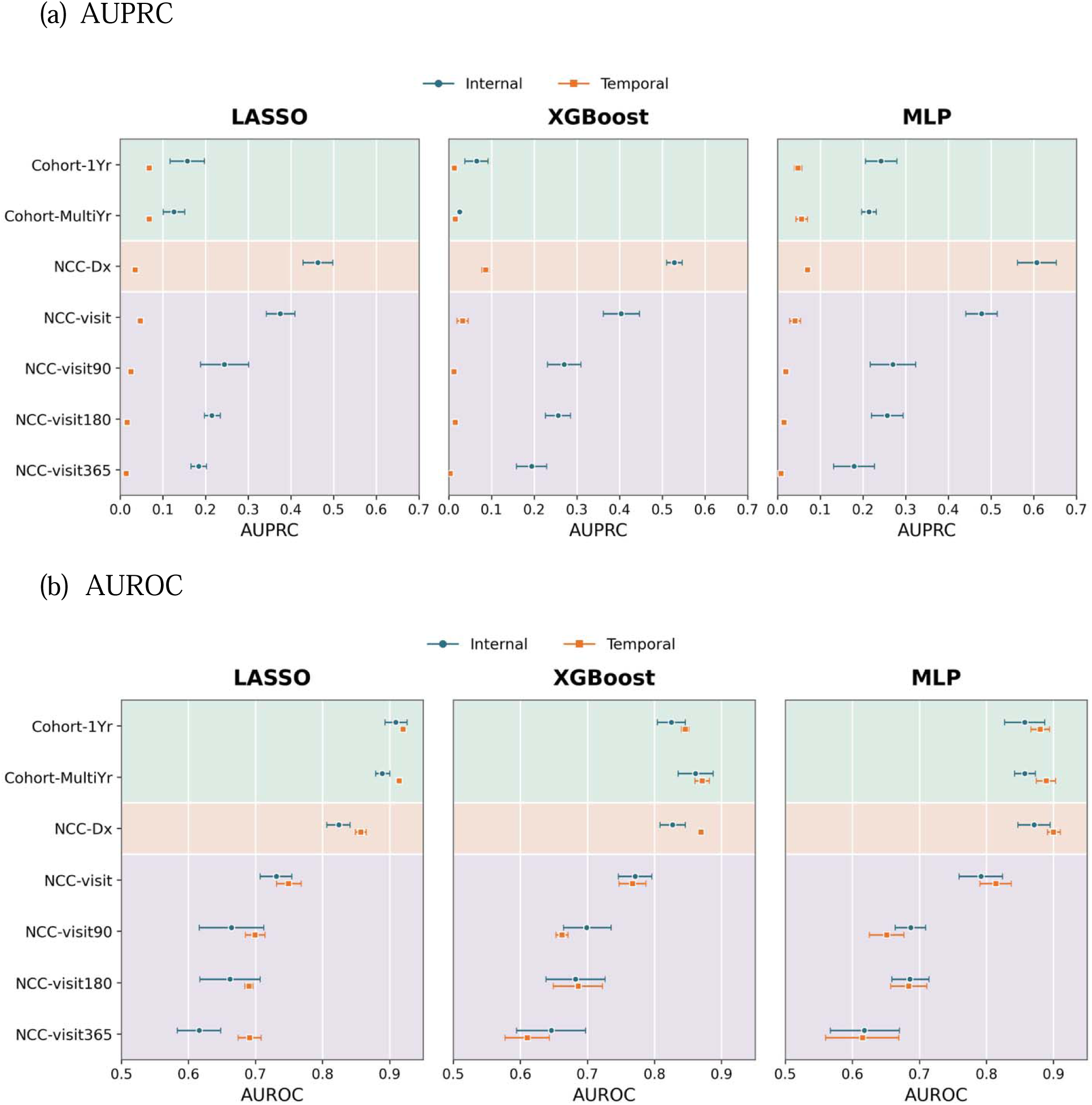
Internal and temporal model discrimination across study designs and models. (a) area under the precision-recall curve (AUPRC) and (B) area under the receiver operating characteristic curve (AUROC) are shown with 95% confidence intervals for LASSO, XGBoost, and multilayer perceptron (MLP) models. Internal estimates were obtained from the held-out outer cross-validation folds. Temporal estimates were obtained by applying each outer-fold model to the temporal evaluation cohort.

We thus focus our evaluation of model discrimination using AUPRC from the temporal cohort. We found that no single study design consistently achieved the highest temporal AUPRC across algorithms. As a baseline, a random model would achieve an AUPRC equal to the prevalence of cases in the sample, which was 0.0012, or 0.12%, in the temporal cohort. For LASSO, the highest temporal AUPRC 0.068 [95% CI, 0.065-0.071], approximately 57 times the baseline. For XGBoost and MLP, the highest temporal AUPRCs were 0.086 (95% CI, 0.077–0.094), approximately 72 times the baseline, and 0.070 (95% CI, 0.065–0.076), approximately 58 times the baseline, respectively. Both were achieved under the NCC design.

### Model calibration

Model calibration was shown in Figure 3. Before recalibration, predicted probabilities were substantially larger than the event rates observed in the temporal cohort for all tested designs. The discrepancy was especially pronounced at the upper end of the predicted-risk distribution: some patient groups received raw predictions approaching 50%-90% even though their observed event rates remained below approximately 1%. This suggests that raw model outputs cannot be interpreted directly as two-year outpatient EOGIC risks.

**Figure 3.**
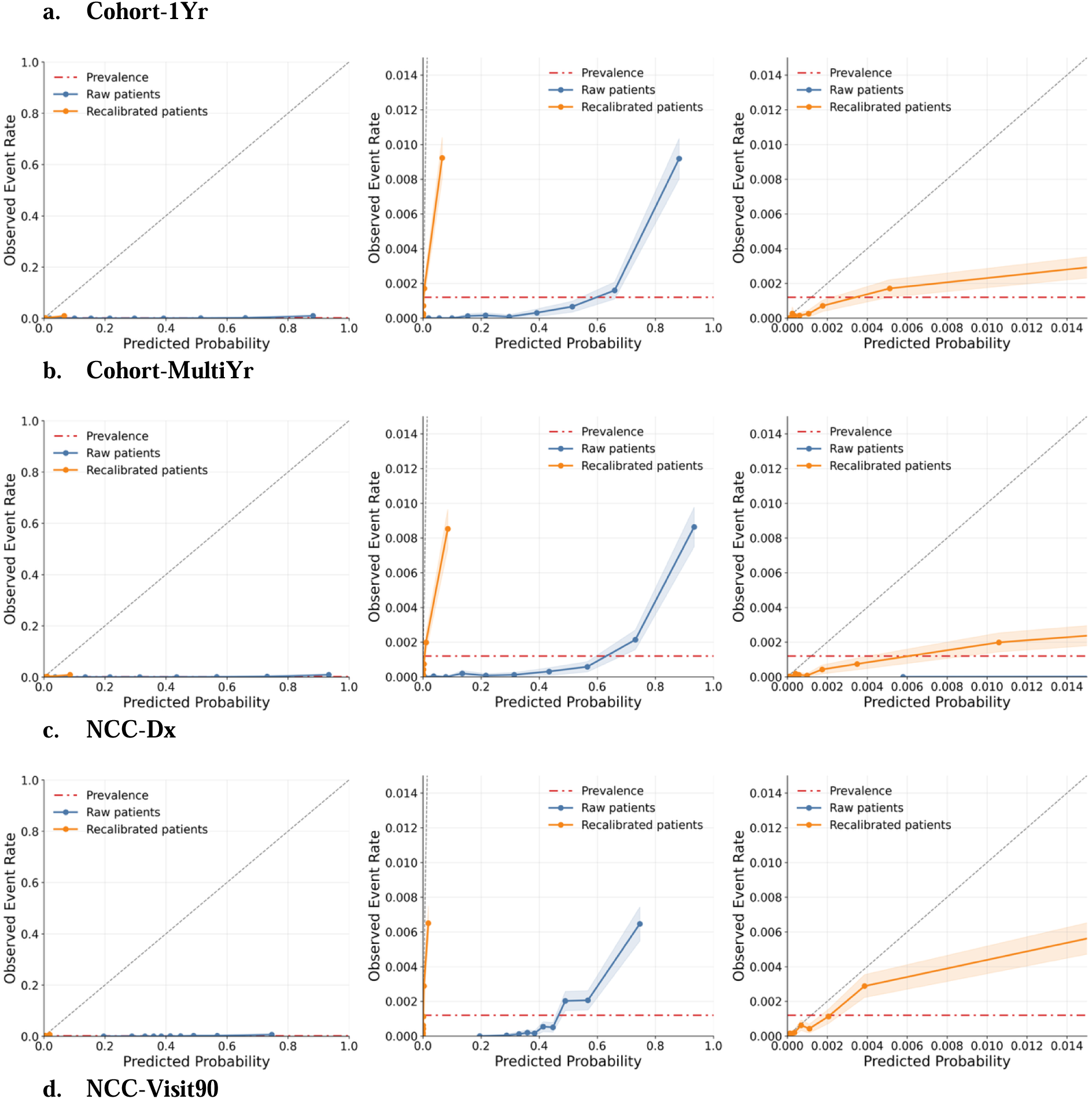
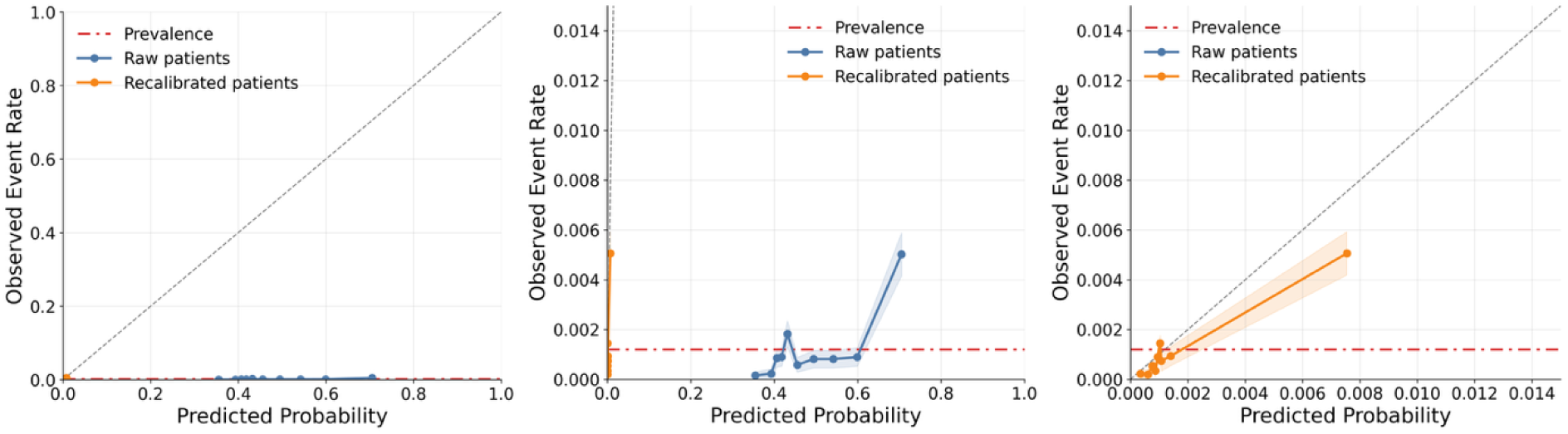
Calibration plots of LASSO model before and after isotonic recalibration in the temporal evaluation cohort. Patients were grouped into deciles of predicted probability; points represent the mean predicted probability and observed EOGIC event rate within each decile. The full probability range (left), an expanded view of the y-axis (middle), and an expanded view of both axes (right) are shown. The diagonal line represents perfect calibration, and the horizontal dashed line represents EOGIC prevalence in the temporal cohort.

Isotonic recalibration compressed predictions into a range more consistent with the approximately 0.12% temporal prevalence and brought predicted and observed risks closer together. However, the enlarged low-risk panels show remaining departures from the 45-degree line, especially in the highest predicted-risk groups. Recalibration therefore corrected much of the gross probability-scale mismatch but did not establish perfect individual-level calibration. Calibration plots for XGBoost and MLP showed the same compression in predicted risk after recalibration (Supplementary Figures 2 and 3).

The change in probability scale was reflected in the Brier scores (Supplementary Table 4). Before recalibration, Brier scores exceeded the value expected for predictions centered near the 0.12% temporal cohort prevalence, indicating poorly scaled predicted probabilities. Isotonic recalibration markedly reduced the Brier scores, with several approaching 0.0012. However, because a model assigning approximately the cohort prevalence to every patient would achieve a similar score, these low post-recalibration values should not directly be interpreted as evidence of accurate risk prediction. Following isotonic recalibration, AUROC and AUPRC were largely preserved across all designs and algorithms with small changes potentially reflecting ties introduced by the non-strictly monotonic isotonic transformation (Supplementary Table 4).

### Feature importance

Figure 4 shows that agreement in learned feature rankings depends strongly on both the modeling algorithm and the study design. Between cohort and NCC designs, feature rankings were more robust under the cohort designs than under the NCC designs; that is, cohort-based models retained more similar feature rankings when the modeling algorithm changed. MLP rankings showed moderate overlap with those from cohort-based XGBoost models but little overlap with those from NCC-based XGBoost models. Similarly, MLP rankings showed modest agreement with cohort-based LASSO rankings and little agreement with most NCC-based LASSO rankings. LASSO and XGBoost also showed some agreement under the two cohort designs, whereas little cross-algorithm agreement was evident among the NCC designs.

**Figure 4.**
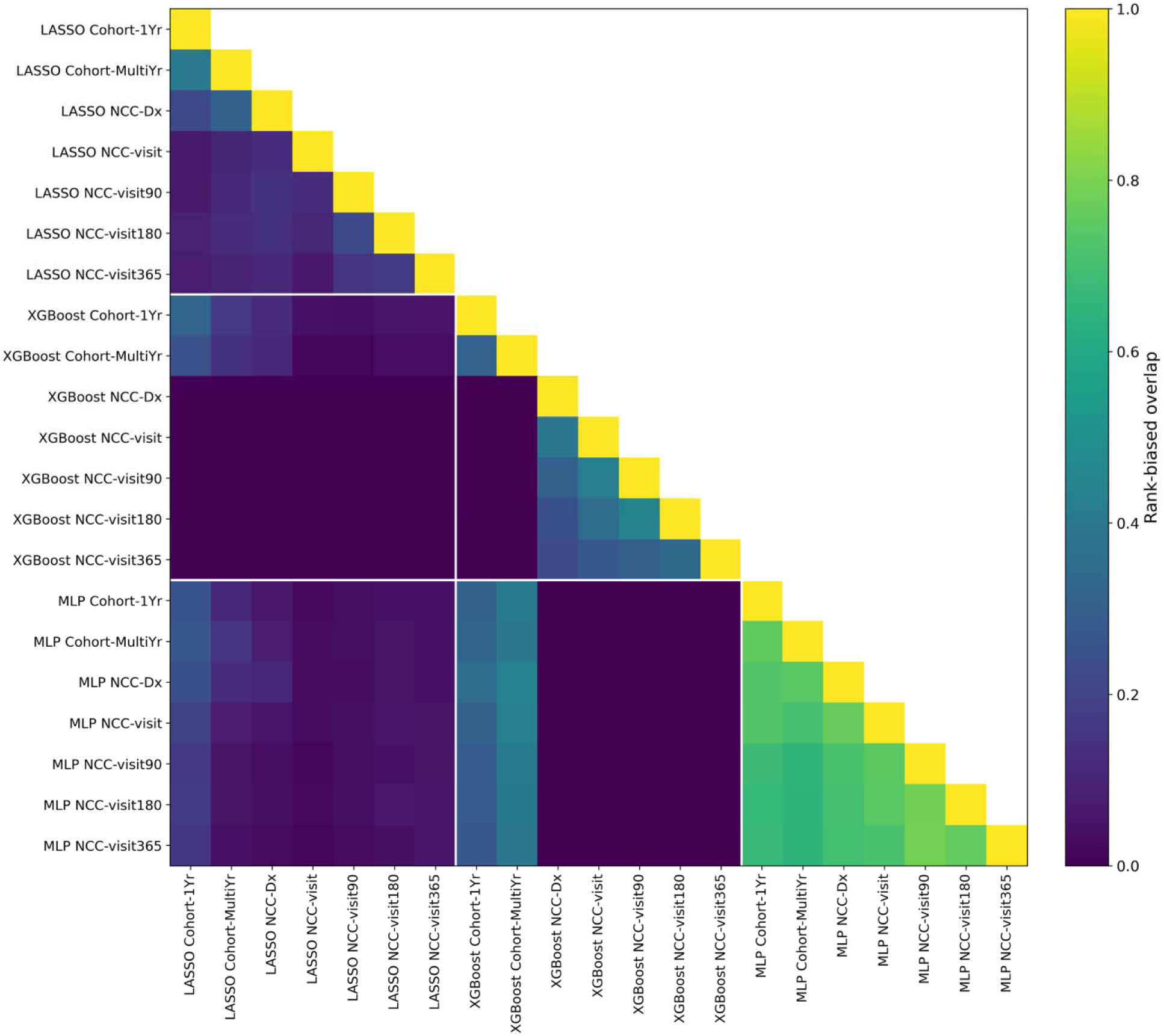
Similarity of feature rankings across modeling approaches and study designs. The heatmap shows pairwise rank-biased overlap (RBO) among feature rankings from all model– design combinations. Higher values indicate greater similarity between feature rankings.

Among the three algorithms, MLP feature rankings appeared least affected by study design, with pairwise RBO values generally in the moderate-to-high range. Despite differences in analytic populations and index-date definitions, MLP retained a comparatively stable hierarchy of important predictors. For XGBoost, the two cohort-based models were moderately similar, whereas the NCC models formed a more internally similar cluster. The limited similarity between cohort-based and NCC-based models suggests that XGBoost feature rankings are sensitive to construction of the analytic dataset. LASSO feature rankings were generally less stable, with the strongest agreement observed between the two cohort designs and some additional agreement with the NCC-Dx-based model. The absence of a cohesive NCC cluster suggests that LASSO rankings were relatively robust across cohort designs but sensitive to index-date anchoring choices within NCC designs.

The magnitudes of feature importance across models and designs are shown in Supplementary Figures 4-7. Although feature importance varied by study design and algorithm, several features recurred among highly ranked predictors across study-design-model combinations, suggesting their potential relevance for EOGIC prediction. CEA measurement recording (mean signed coefficient, +2.201), anorectal disorders (+1.744), pancreatic disorders (+1.652), rectal or anal hemorrhage (+1.630), and intestinal disorders (+1.482) were prominent positive predictors based on LASSO model coefficients. These features also had relatively high feature importance in XGBoost and MLP models based on their corresponding feature importance measure.

## DISCUSSION

In this study, we showed that the choice of study design substantially influenced model discrimination, calibration, and feature-ranking patterns of models for EOGIC prediction. One discrimination metric, AUPRC, is particularly sensitive to the outcome prevalence; therefore, model performance evaluated in case-control samples did not translate to performance in the intended clinical population, where EOGIC prevalence is much lower. All models, including those developed using cohort designs, substantially overestimated the EOGIC risk. This finding suggests that recalibrating EOGIC prediction models is essential for aligning predicted risks with absolute risk scale.

Our findings extend prior work showing that case-control sampling does not use control samples efficiently and introduces many design decisions that may be difficult to reproduce. Case-control designs are traditionally favored when outcomes are rare and collecting or adjudicating predictors for an entire cohort is costly.^33,34^ In the setting of EHR-based risk modeling, however, the data have already been collected, requiring no new data collection. By discarding eligible controls, NCC designs do not fully use information from the broader population of patients who do not experience the outcome.^19,35^ Although NCC designs can reduce computational burden and facilitate rapid model development, they change outcome prevalence and require decisions about matching, case-to-control ratios, control reuse, and control index dates. These decisions are often insufficiently reported, which can make NCC designs challenging to reproduce.

Even when a case-control design is well specified and the results are internally valid, AUPRC estimated in its case-enriched sample does not directly represent performance in the intended clinical population. Because AUPRC depends on outcome prevalence, it must be evaluated in a sample with prevalence representative of the intended deployment setting to support clinically meaningful interpretation. While AUROC does not depend on outcome prevalence, it is much less sensitive compared to AUPRC. In our study, AUPRC’s expected value under random classification equals the outcome prevalence itself (∼0.12% in the temporal cohort); on this scale, the observed temporal AUPRC values corresponded to roughly 2.5-fold to 72-fold enrichment over the prevalence baseline across all models. In contrast, AUROC’s expected value under random classification is fixed at 0.5, the observed temporal AUROC values corresponded to only approximately 1.2-fold to 1.8-fold enrichment above chance between the best– and worst-performing models. Thus, although both metrics are useful for comparing prediction models, AUPRC may be more informative for evaluating discrimination in early-onset cancers.

Calibration was another central consideration. Our findings are consistent with prior evidence that outcome enrichment under NCC design can shift predicted probabilities away from the absolute risks.^36^ We further showed that when the outcome is rare, such as EOGIC, models developed under cohort design also substantially overestimated the absolute risk of EOGIC. This finding highlights that model recalibration should therefore be considered regardless of study design to better align predicted probabilities with observed risk in the intended clinical population.

This study has several limitations. First, all data were obtained from a single regional health system; therefore, we did not assess how study design choices affect model transportability across institutions with different patient populations, healthcare practices, and EHR documentation patterns. Second, we could not vary every possible design decision within an NCC study, such as matching specifications, control reuse, and case-to-control ratios. Third, we compared LASSO, XGBoost, and MLP models but did not evaluate Transformer-based risk prediction models; thus, the generalizability of these findings to Transformer-based approaches remains unknown. Finally, we did not evaluate how study design choices affect benefit-risk tradeoffs across clinically relevant decision thresholds. Such analyses, together with prospective evaluation, are needed to determine the clinical consequences of using these models to guide early detection at the point of care.

## CONCLUSION

Study design substantially affects the discrimination, calibration, and feature-ranking patterns of EHR-based EOGIC prediction models. Cohort designs are generally better suited to EHR-based models for early detection because they preserve the outcome incidence and temporal relationship between the predictors and the outcome. When case-control designs are used for model development, models should be evaluated and recalibrated in an incidence-representative cohort that reflects the intended clinical setting.

### Data Availability

The EHR data used in this study contain protected health information and are not publicly available. Aggregated data to support the findings of this study can be made available upon a reasonable request from the corresponding author.

### Code Availability

Code will be made publicly available at https://github.com/causailab/study_design_comparison_rare_outcomes

## Acknowledgments

This work was delivered as part of the PROSPECT team supported by the Cancer Grand Challenges partnership funded by Cancer Research UK (CGCATF-2023/100037 to Y.C.) and the National Cancer Institute (OT2CA297576 to Y.C.). The funders had no role in study design, data collection and analysis, decision to publish or preparation of the manuscript.

## Author contributions

L.Z. and R.F. had full access to all the data in the study and take responsibility for the integrity of the data and the accuracy of the data analysis. R.F., Y.C., and L.Z. conceived and designed the study. R.F. and L.Z. performed the statistical analysis. All authors contributed to the interpretation of data. R.F. and L.Z. drafted the manuscript. All authors critically revised the manuscript for important intellectual content and approved the final version of the manuscript.

