## Supplementary Information for "Cohort Versus Nested Case-Control Designs for Predicting Early-Onset Gastrointestinal Cancer Using Electronic Health Records"

**Cohort Versus Nested Case-Control Design for Prediction Models of Rare Outcomes Using Electronic Health Records**

**Supplementary Materials**

**Supplementary Table 1. Study design definitions and analytic sample sizes.**

| **Study design** | **Design family** | **Index date definition** | **Cases (n)** | **Controls/noncases (n)** | **Predictor window** | **Outcome window** |
| --- | --- | --- | --- | --- | --- | --- |
| Cohort-1Yr | Cohort | First qualifying outpatient visit in 2022 | 303 | 256,551 noncases | 365 days before index | Incident EOGIC within 2 years |
| Cohort-MultiYr | Cohort | First qualifying outpatient visit during 2020-2022 | 711 | 414,818 noncases | 365 days before index | Incident EOGIC within 2 years |
| NCC-Dx | Nested case-control | Cancer diagnosis date for cases; matched outpatient visit date for controls | 711 | 1:10 matched controls | 365 days before index | Case diagnosis anchor; controls event-free |
| NCC-visit | Nested case-control | Outpatient visit before and closest to diagnosis | 352 | 1:10 matched controls | 365 days before index | Case diagnosis anchor; controls event-free |
| NCC-visit90 | Nested case-control | Outpatient visit at least 90 days before diagnosis | 257 | 1:10 matched controls | 365 days before index | Case diagnosis anchor; controls event-free |
| NCC-visit180 | Nested case-control | Outpatient visit at least 180 days before diagnosis | 245 | 1:10 matched controls | 365 days before index | Case diagnosis anchor; controls event-free |
| NCC-visit365 | Nested case-control | Outpatient visit at least 365 days before diagnosis | 209 | 1:10 matched controls | 365 days before index | Case diagnosis anchor; controls event-free |

**Supplementary Table 2. Internal and temporal discrimination across study designs and modeling approaches.** Values are mean AUROC or AUPRC across five outer folds with 95% confidence intervals. Temporal metrics summarize performance when each outer-fold model was applied independently to the temporal cohort.

| **Study design** | **Model** | **Internal AUROC** | **Internal AUPRC** | **Temporal AUROC** | **Temporal AUPRC** |
| --- | --- | --- | --- | --- | --- |
| Cohort-1Yr | LASSO | **0.909 (0.893-0.926)** | 0.157 (0.117-0.197) | **0.920 (0.916-0.923)** | 0.068 (0.065-0.072) |
| Cohort-1Yr | XGBoost | 0.825 (0.804-0.846) | 0.065 (0.037-0.092) | 0.846 (0.840-0.851) | 0.013 (0.010-0.016) |
| Cohort-1Yr | MLP | 0.857 (0.827-0.887) | 0.242 (0.206-0.279) | 0.880 (0.866-0.894) | 0.048 (0.039-0.057) |
| Cohort-MultiYr | LASSO | 0.889 (0.879-0.900) | 0.126 (0.101-0.151) | 0.914 (0.910-0.917) | 0.068 (0.065-0.071) |
| Cohort-MultiYr | XGBoost | 0.861 (0.835-0.887) | 0.025 (0.022-0.028) | 0.871 (0.860-0.882) | 0.015 (0.014-0.015) |
| Cohort-MultiYr | MLP | 0.857 (0.842-0.873) | 0.214 (0.197-0.231) | 0.889 (0.874-0.903) | 0.056 (0.043-0.070) |
| NCC-Dx | LASSO | 0.824 (0.806-0.841) | 0.463 (0.428-0.498) | 0.857 (0.849-0.865) | 0.035 (0.032-0.037) |
| NCC-Dx | XGBoost | 0.827 (0.808-0.846) | 0.528 (0.510-0.546) | 0.869 (0.865-0.873) | **0.086 (0.077-0.094)** |
| NCC-Dx | MLP | 0.871 (0.847-0.895) | **0.607 (0.562-0.653)** | 0.900 (0.891-0.910) | 0.070 (0.065-0.076) |
| NCC-visit | LASSO | 0.731 (0.707-0.754) | 0.375 (0.342-0.409) | 0.749 (0.731-0.768) | 0.047 (0.044-0.050) |
| NCC-visit | XGBoost | 0.771 (0.746-0.796) | 0.404 (0.362-0.446) | 0.767 (0.747-0.787) | 0.032 (0.019-0.045) |
| NCC-visit | MLP | 0.792 (0.759-0.824) | 0.478 (0.441-0.514) | 0.814 (0.790-0.837) | 0.041 (0.028-0.054) |
| NCC-visit90 | LASSO | 0.664 (0.616-0.712) | 0.244 (0.188-0.301) | 0.699 (0.685-0.714) | 0.025 (0.020-0.030) |
| NCC-visit90 | XGBoost | 0.699 (0.664-0.735) | 0.270 (0.231-0.309) | 0.662 (0.653-0.671) | 0.012 (0.008-0.017) |
| NCC-visit90 | MLP | 0.687 (0.664-0.709) | 0.270 (0.217-0.323) | 0.651 (0.625-0.677) | 0.019 (0.013-0.025) |
| NCC-visit180 | LASSO | 0.662 (0.617-0.707) | 0.215 (0.197-0.234) | 0.690 (0.684-0.696) | 0.016 (0.012-0.020) |
| NCC-visit180 | XGBoost | 0.682 (0.638-0.726) | 0.256 (0.226-0.285) | 0.686 (0.649-0.722) | 0.015 (0.010-0.019) |
| NCC-visit180 | MLP | 0.686 (0.659-0.714) | 0.257 (0.220-0.294) | 0.684 (0.657-0.711) | 0.015 (0.012-0.018) |
| NCC-visit365 | LASSO | 0.616 (0.583-0.648) | 0.184 (0.166-0.202) | 0.691 (0.674-0.708) | 0.014 (0.012-0.016) |
| NCC-visit365 | XGBoost | 0.646 (0.594-0.697) | 0.194 (0.158-0.229) | 0.610 (0.577-0.643) | 0.003 (0.002-0.003) |
| NCC-visit365 | MLP | 0.618 (0.567-0.670) | 0.179 (0.131-0.227) | 0.615 (0.560-0.669) | 0.008 (0.002-0.013) |

**Supplementary Table 3** Hyperparameter search space for MLP models

(a) Hyperparameter search space and fixed training parameters

| **Parameter** | **Value(s)** |
| --- | --- |
| Hidden-layer architecture | 128; 128-32; 256-64; 256-128-32 |
| Dropout rate | 0.1; 0.3 |
| Learning rate | 0.001; 0.0003 |
| Weight decay | 0.0001 |
| Positive-class weight | Noncase-to-case ratio within each training subset |
| Optimizer | AdamW |
| Loss function | Weighted binary cross-entropy |
| Hidden-layer activation | ReLU |
| Batch normalization | Applied after each hidden linear layer |
| Training batch size | 1,024 |
| Evaluation batch size | 4,096 |
| Maximum epochs | 50 |
| Early-stopping patience | 5 epochs |
| Gradient clipping | Maximum norm of 5.0 |
| Outer cross-validation folds | 5 |
| Inner cross-validation folds | 3 |
| Hyperparameter-selection metric | Mean inner-validation AUPRC |

(b) selected MLP configuration by study design

| **Study design** | **Hidden layers** | **Dropout** | **Learning rate** | **Weight decay** | **Class weighting** | **Selected in folds** |
| --- | --- | --- | --- | --- | --- | --- |
| Cohort-1Yr | 256-128-32 | 0.3 | 0.0003 | 0.0001 | balanced | 3/5 |
| Cohort-MultiYr | 128-32 | 0.3 | 0.0003 | 0.0001 | balanced | 3/5 |
| NCC-Dx | 256-64 | 0.3 | 0.0003 | 0.0001 | balanced | 2/5 |
| NCC-visit | 256-64 | 0.3 | 0.0003 | 0.0001 | balanced | 2/5 |
| NCC-visit90 | no dominant set | mixed | mixed | 0.0001 | balanced | 1/5 each |
| NCC-visit180 | no dominant set | mixed | mixed | 0.0001 | balanced | 1/5 each |
| NCC-visit365 | no dominant set | mixed | mixed | 0.0001 | balanced | 1/5 each |

**Supplementary Figure 1. Internal and temporal precision–recall curves across study designs. Precision-Recall curves are shown for (a) LASSO, (b) XGBoost, and (c) MLP. The left panel shows internal evaluation and the right panel shows the temporal evaluation.**

(a) LASSO

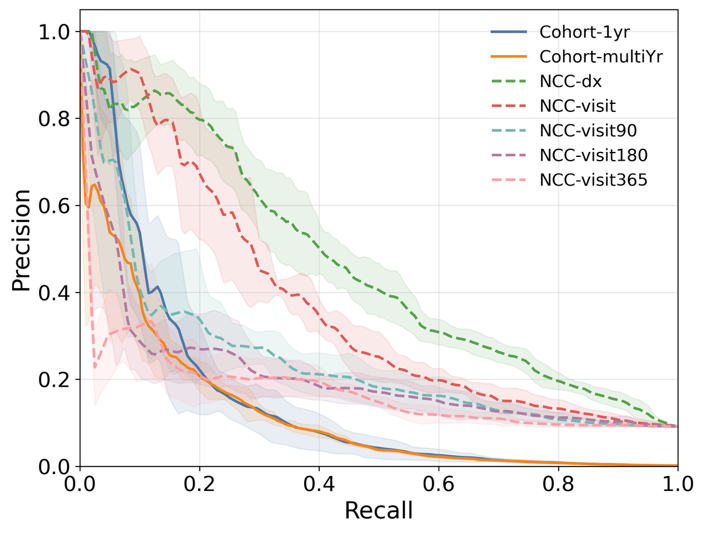

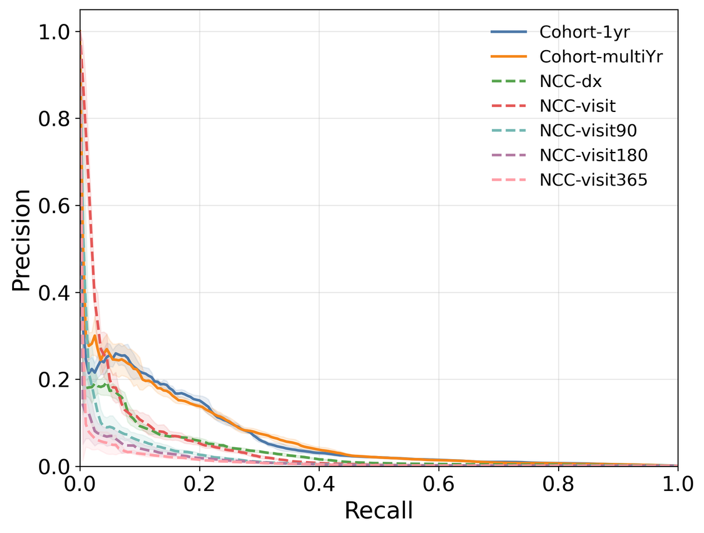

(b) XGBoost

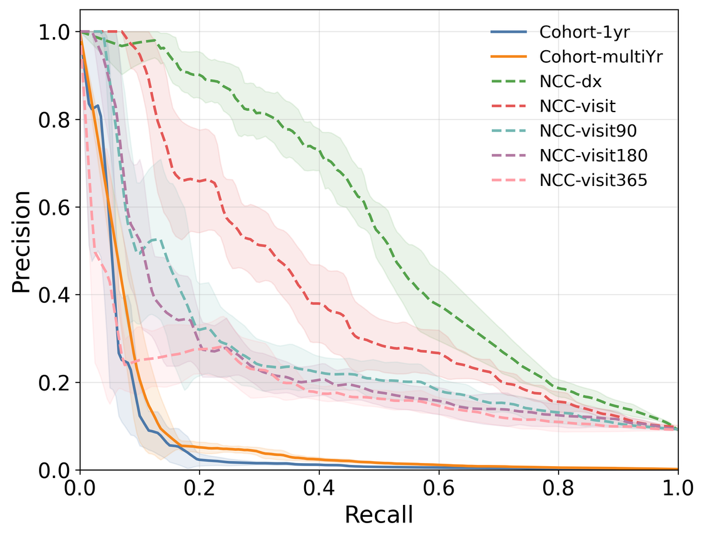

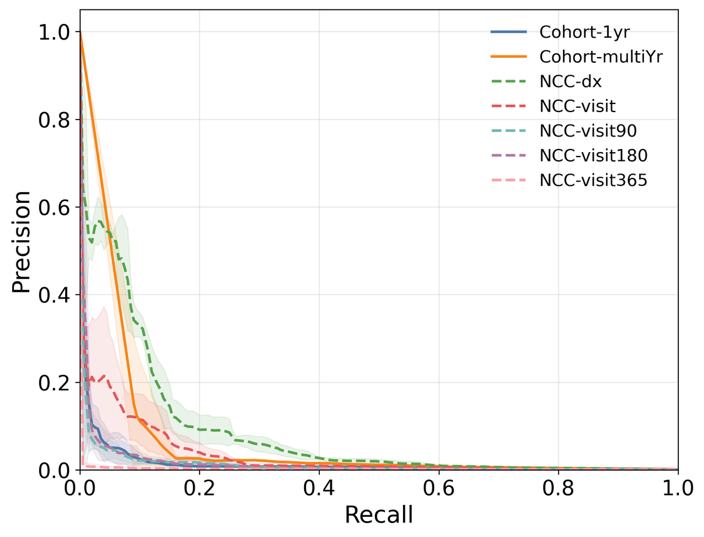

(c) MLP

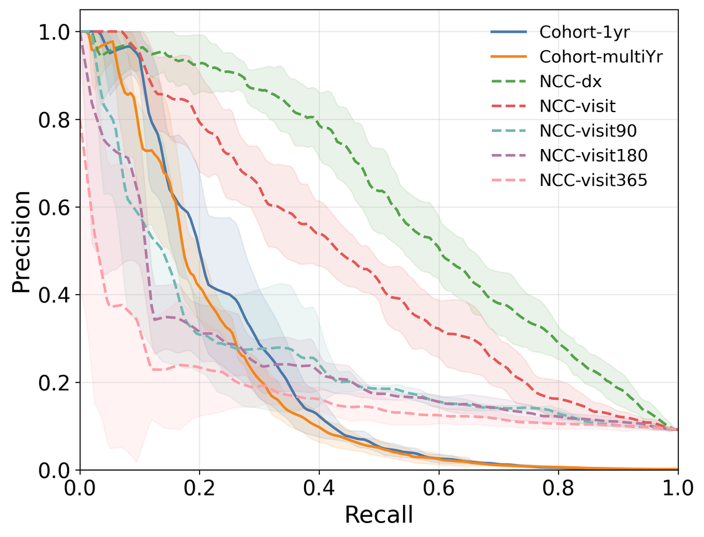

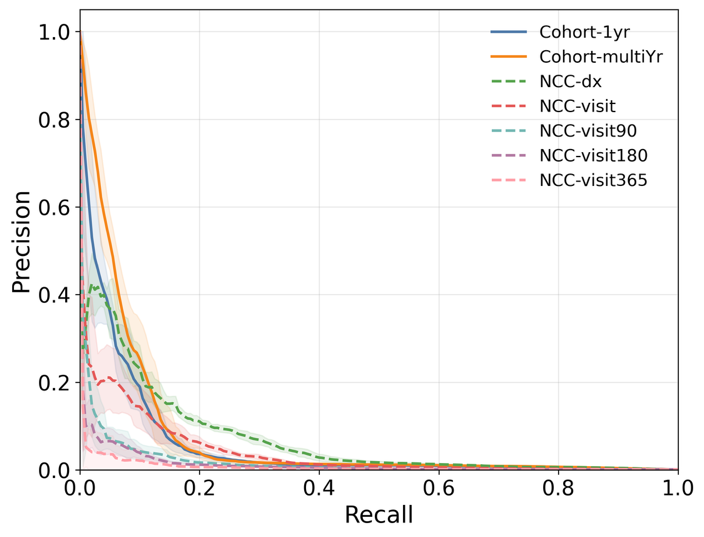

**Supplementary Figure 2. Calibration plots for XGBoost before and after isotonic recalibration in the temporal cohort.** Patients were grouped into deciles of predicted probability. Points represent the mean predicted probability and observed EOGIC event rate within each decile. The diagonal line represents perfect calibration, and the horizontal dashed line represents EOGIC cumulative incidence in the temporal cohort.

(a) Cohort-1Yr

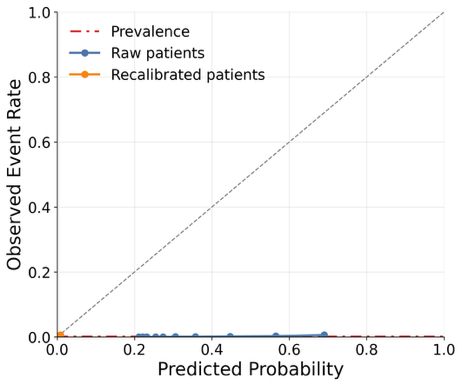

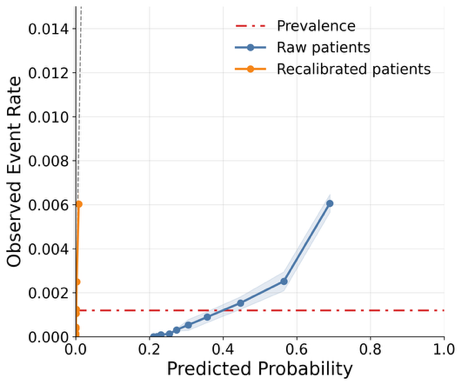

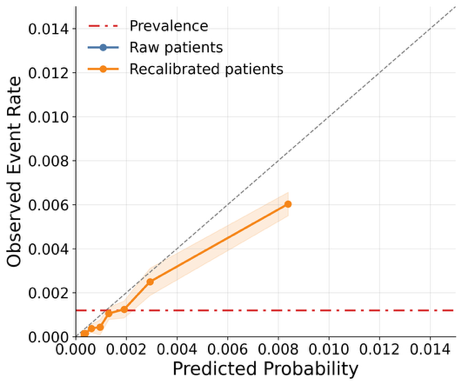

(b) Cohort-MultiYr

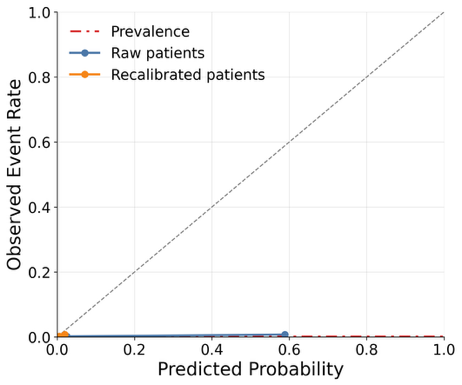

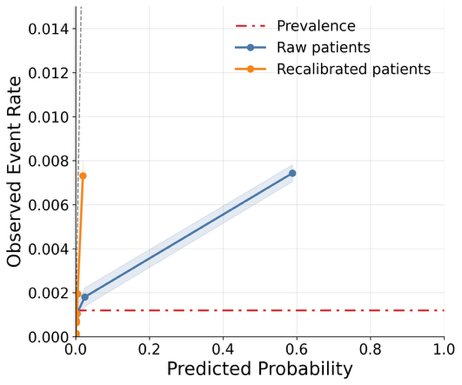

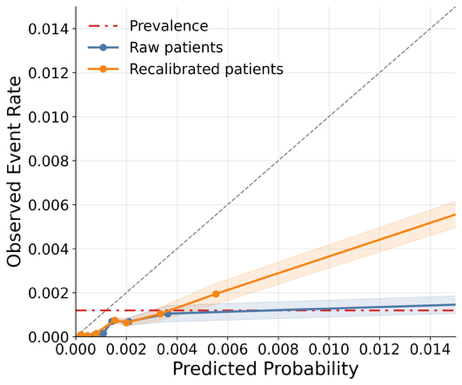

(c) NCC-Dx

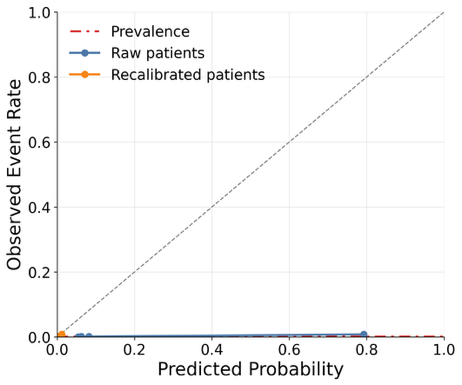

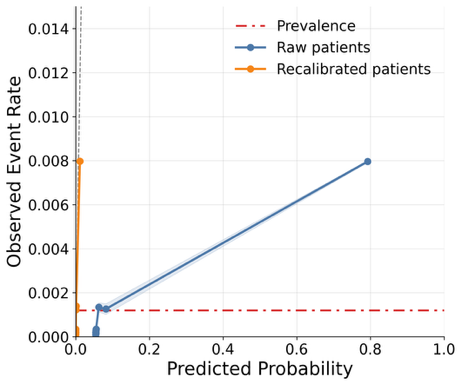

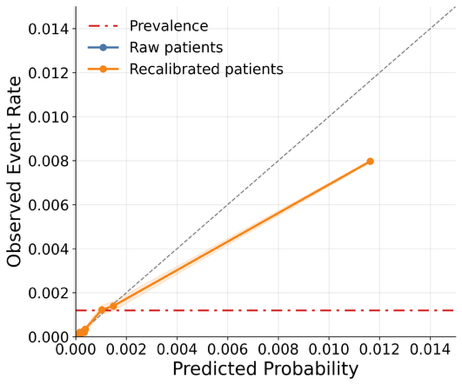

(d) NCC-Visit90

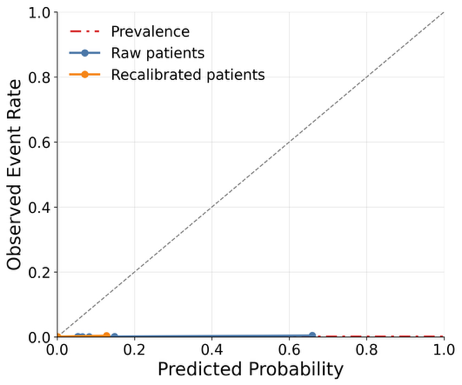

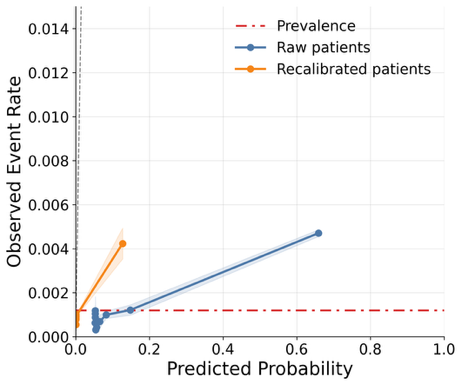

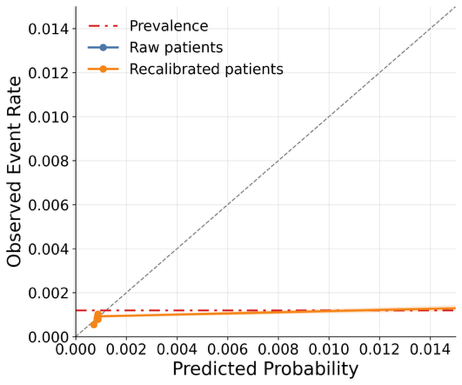

**Supplementary Figure 3. Calibration plots for MLP before and after isotonic recalibration in the temporal cohort across study designs.** Patients were grouped into deciles of predicted probability. Points represent the mean predicted probability and observed EOGIC event rate within each decile. The diagonal line represents perfect calibration, and the horizontal dashed line represents EOGIC cumulative incidence in the temporal cohort.

(a) Cohort-1Yr

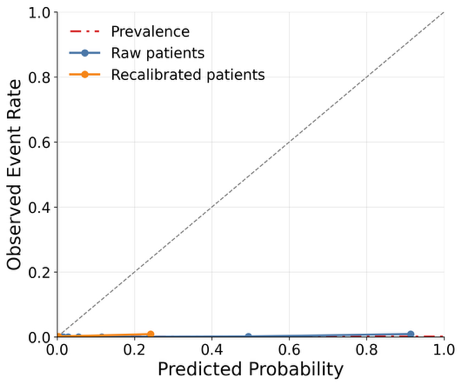

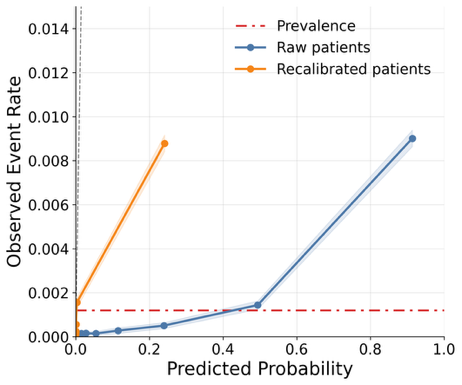

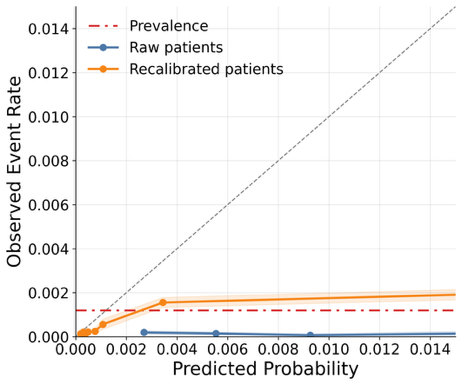

(b) Cohort-MultiYr

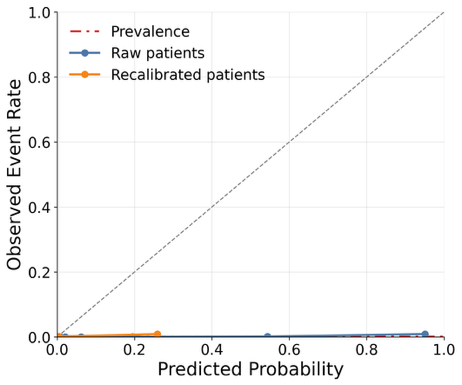

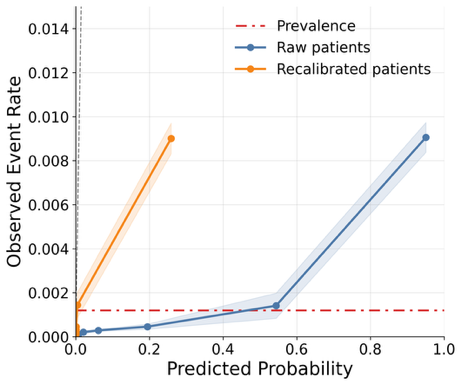

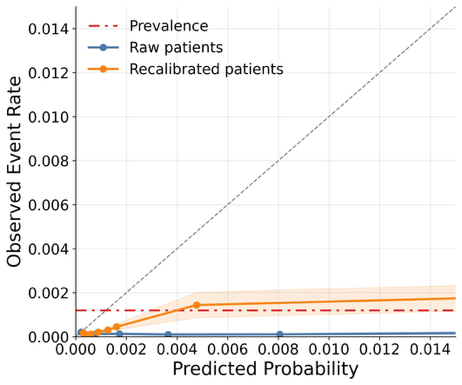

(c) NCC-Dx

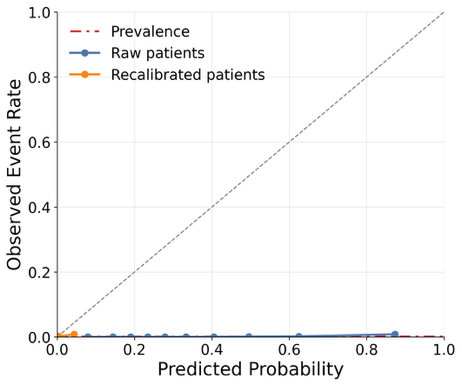

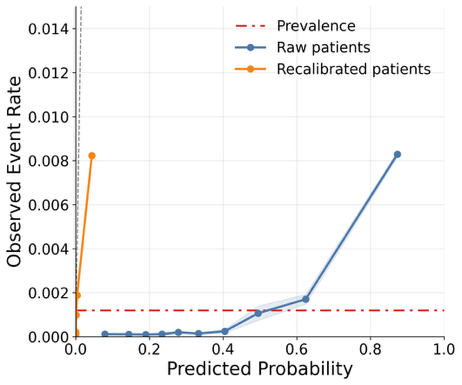

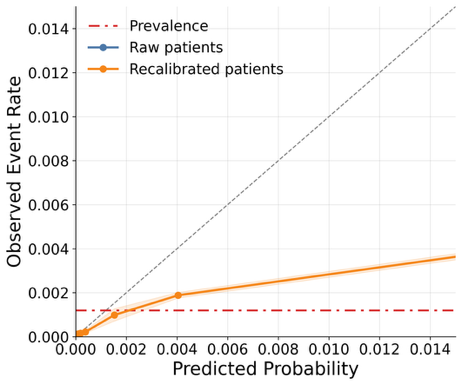

(d) NCC-Visit90

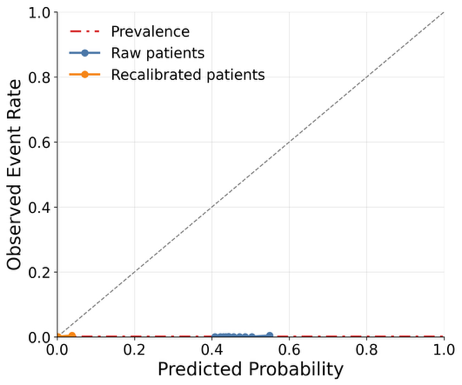

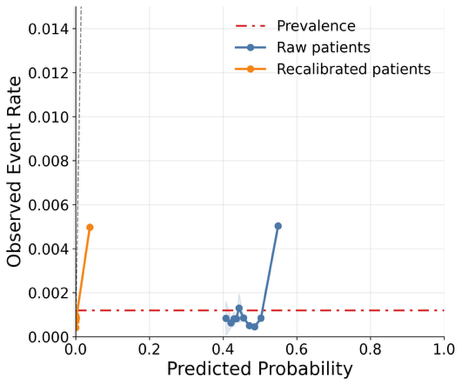

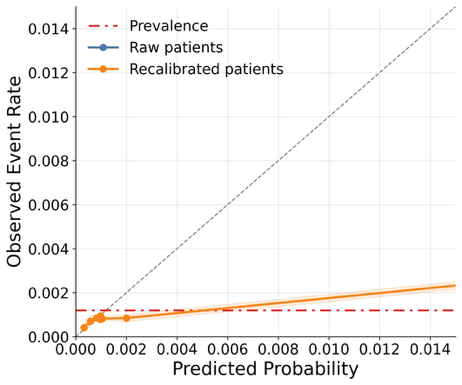

**Supplementary Table 4. Discrimination and Brier scores before and after isotonic recalibration in the temporal cohort.** Results are shown for (A) LASSO, (B) XGBoost, and (C) multilayer perceptron (MLP) models. Before-recalibration values were calculated from the original model predictions, and after-recalibration values were calculated following isotonic recalibration. Lower Brier scores indicate better calibration.

(a) LASSO

| **Study design** | **Before calibration** | | | **After calibration** | | |
| --- | --- | --- | --- | --- | --- | --- |
|  | **AUROC** | **AUPRC** | **Brier score** | **AUROC** | **AUPRC** | **Brier score** |
| Cohort-1Yr | **0.920** | **0.068** | **0.1806** | **0.9227** | 0.0653 | 0.0023 |
| Cohort-MultiYr | 0.914 | **0.068** | 0.2088 | 0.9161 | **0.0730** | 0.0023 |
| NCC-Dx | 0.857 | 0.0347 | 0.2005 | 0.8644 | 0.0370 | **0.0012** |
| NCC-visit | 0.749 | 0.0471 | 0.2284 | 0.7816 | 0.0558 | **0.0012** |
| NCC-visit90 | 0.699 | 0.0246 | 0.2411 | 0.7393 | 0.0243 | **0.0012** |
| NCC-visit180 | 0.690 | 0.0164 | 0.2495 | 0.7209 | 0.0204 | **0.0012** |
| NCC-visit365 | 0.691 | 0.0140 | 0.2489 | 0.7413 | 0.0158 | **0.0012** |

(b) XGBoost

| **Study design** | **Before calibration** | | | **After calibration** | | |
| --- | --- | --- | --- | --- | --- | --- |
|  | **AUROC** | **AUPRC** | **Brier score** | **AUROC** | **AUPRC** | **Brier score** |
| Cohort-1Yr | 0.846 | 0.013 | 0.1491 | 0.8597 | 0.0162 | **0.0012** |
| Cohort-MultiYr | **0.871** | 0.015 | 0.0457 | **0.8806** | 0.0160 | **0.0012** |
| NCC-Dx | 0.869 | **0.086** | 0.0700 | 0.8759 | **0.0977** | **0.0012** |
| NCC-visit | 0.767 | 0.032 | 0.0671 | 0.8048 | 0.0227 | 0.0134 |
| NCC-visit90 | 0.662 | 0.012 | 0.0507 | 0.7008 | 0.0103 | 0.0041 |
| NCC-visit180 | 0.686 | 0.015 | 0.0611 | 0.7531 | 0.0200 | **0.0012** |
| NCC-visit365 | 0.610 | 0.003 | **0.0339** | 0.6274 | 0.0023 | **0.0012** |

(c) MLP

| **Study design** | **Before calibration** | | | **After calibration** | | |
| --- | --- | --- | --- | --- | --- | --- |
|  | **AUROC** | **AUPRC** | **Brier score** | **AUROC** | **AUPRC** | **Brier score** |
| Cohort-1Yr | 0.880 | 0.048 | **0.1444** | 0.8751 | 0.0121 | 0.0336 |
| Cohort-MultiYr | 0.889 | 0.056 | 0.1700 | 0.8879 | 0.0154 | 0.0222 |
| NCC-Dx | **0.900** | **0.070** | 0.2381 | **0.9014** | **0.0558** | 0.0016 |
| NCC-visit | 0.814 | 0.041 | 0.2755 | 0.8172 | 0.0329 | 0.0014 |
| NCC-visit90 | 0.651 | 0.019 | 0.2312 | 0.6794 | 0.0144 | 0.0018 |
| NCC-visit180 | 0.684 | 0.015 | 0.2260 | 0.6959 | 0.0116 | 0.0016 |
| NCC-visit365 | 0.615 | 0.008 | 0.2560 | 0.6293 | 0.0060 | **0.0013** |

**Supplementary Table 5. Codeset for EOGIC.**

| Concept Id | Concept Code | Concept Name | Domain | Standard Concept Caption | Descendants |
| --- | --- | --- | --- | --- | --- |
| 196044 | 372014001 | Primary malignant neoplasm of stomach | Condition | Standard | Yes |
| 192836 | 94048009 | Primary malignant neoplasm of small intestine | Condition | Standard | Yes |
| 74582 | 93984006 | Primary malignant neoplasm of rectum | Condition | Standard | Yes |
| 438699 | 93980002 | Primary malignant neoplasm of rectosigmoid junction | Condition | Standard | Yes |
| 199754 | 372003004 | Primary malignant neoplasm of pancreas | Condition | Standard | Yes |
| 201519 | 95214007 | Primary malignant neoplasm of liver | Condition | Standard | Yes |
| 197806 | 372139008 | Primary malignant neoplasm of gallbladder | Condition | Standard | Yes |
| 26638 | 371984007 | Primary malignant neoplasm of esophagus | Condition | Standard | Yes |
| 197500 | 93761005 | Primary malignant neoplasm of colon | Condition | Standard | Yes |
| 194589 | 371970002 | Primary malignant neoplasm of biliary tract | Condition | Standard | Yes |
| 436348 | 93669004 | Primary malignant neoplasm of anal canal | Condition | Standard | Yes |

**Supplementary Figure 4. Top 30 LASSO features ranked by coefficient magnitude in Cohort-MultiYr.** Corresponding coefficients are shown for Cohort-1Yr, NCC-Dx, NCC-Visit, and NCC-visit90.

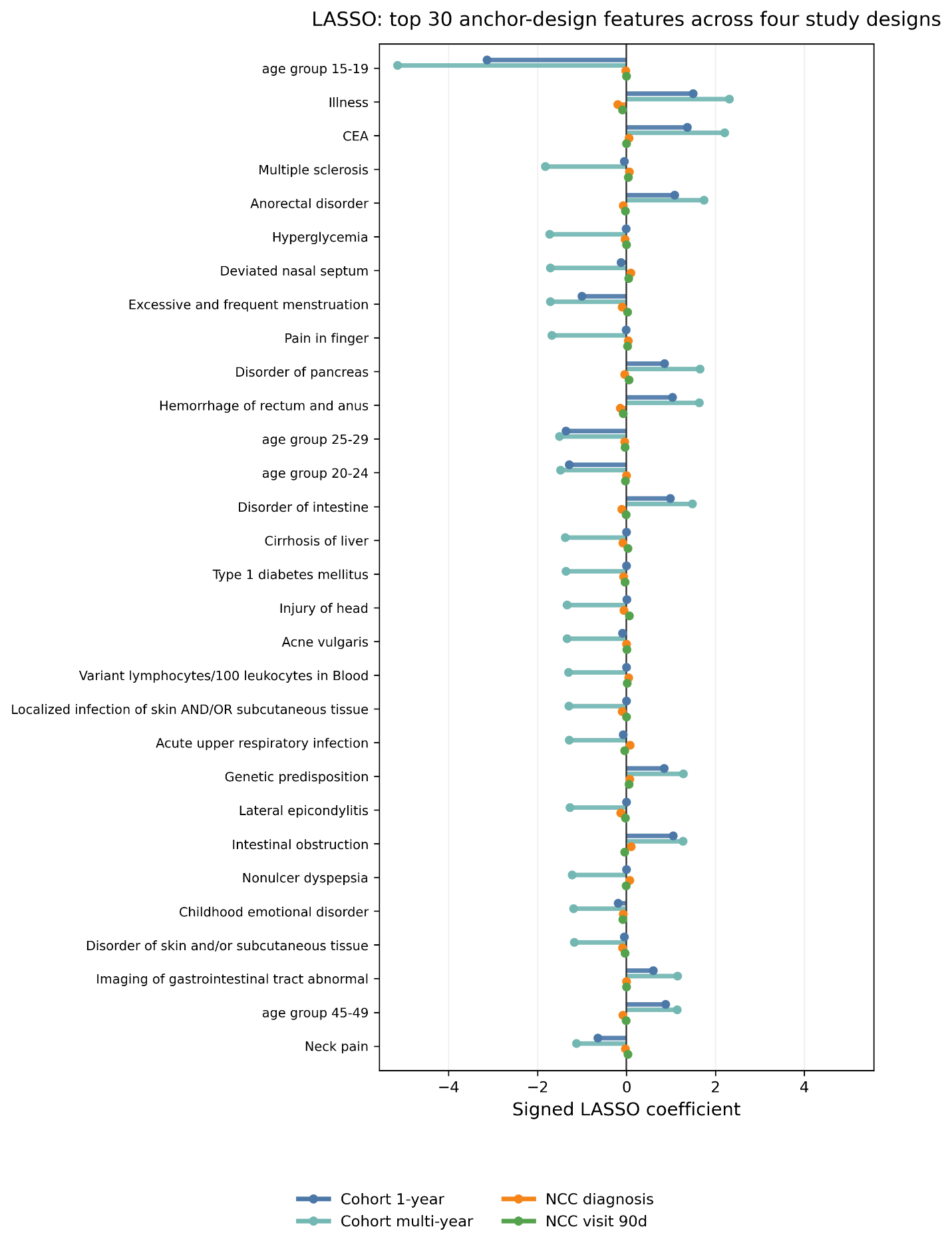

**Supplementary Figure 5. Top 30 LASSO features ranked by coefficient magnitude in NCC-Dx.**

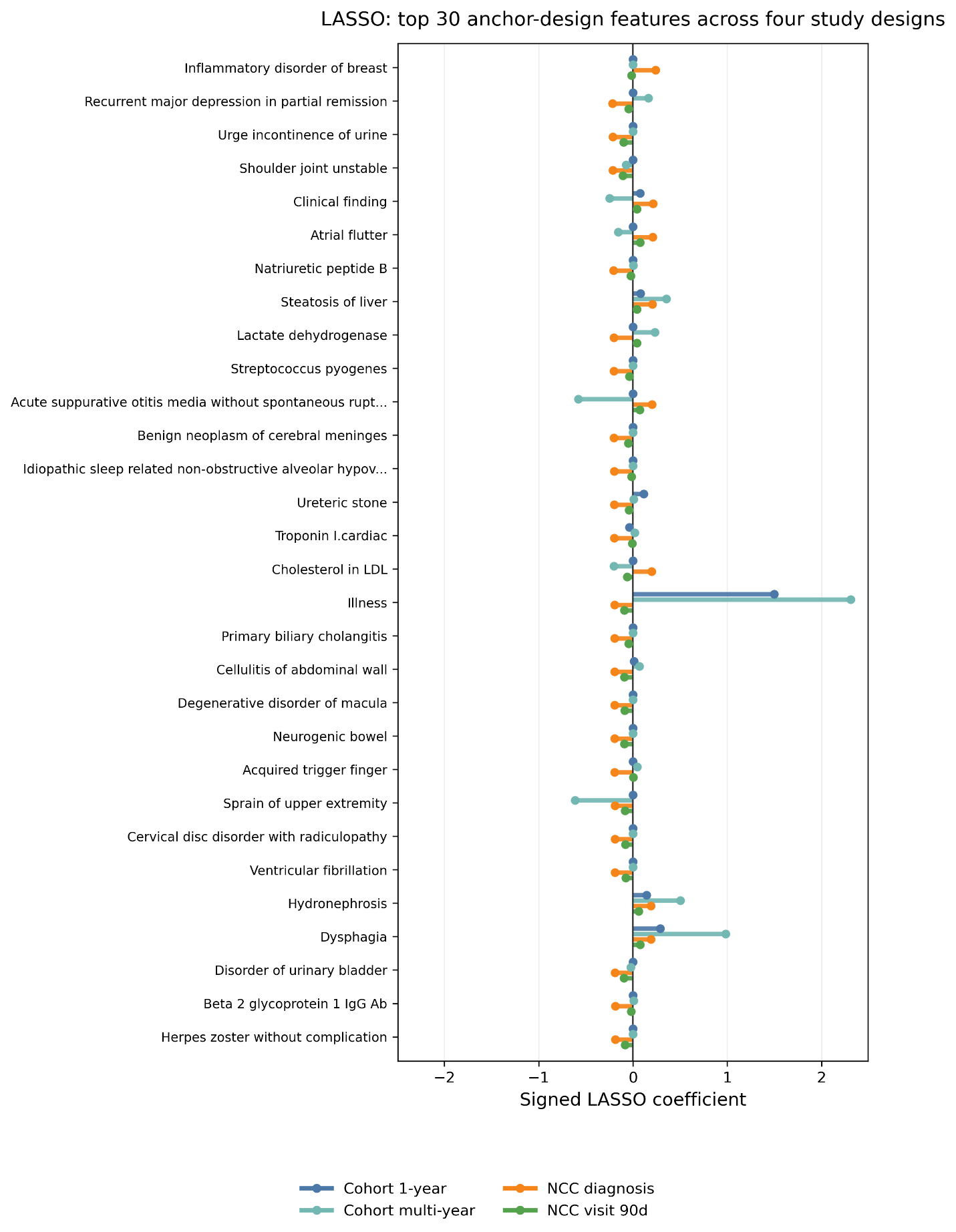

**Supplementary Figure 6. Top 30 XGBoost features ranked by feature importance in Cohort-MultiYr.**

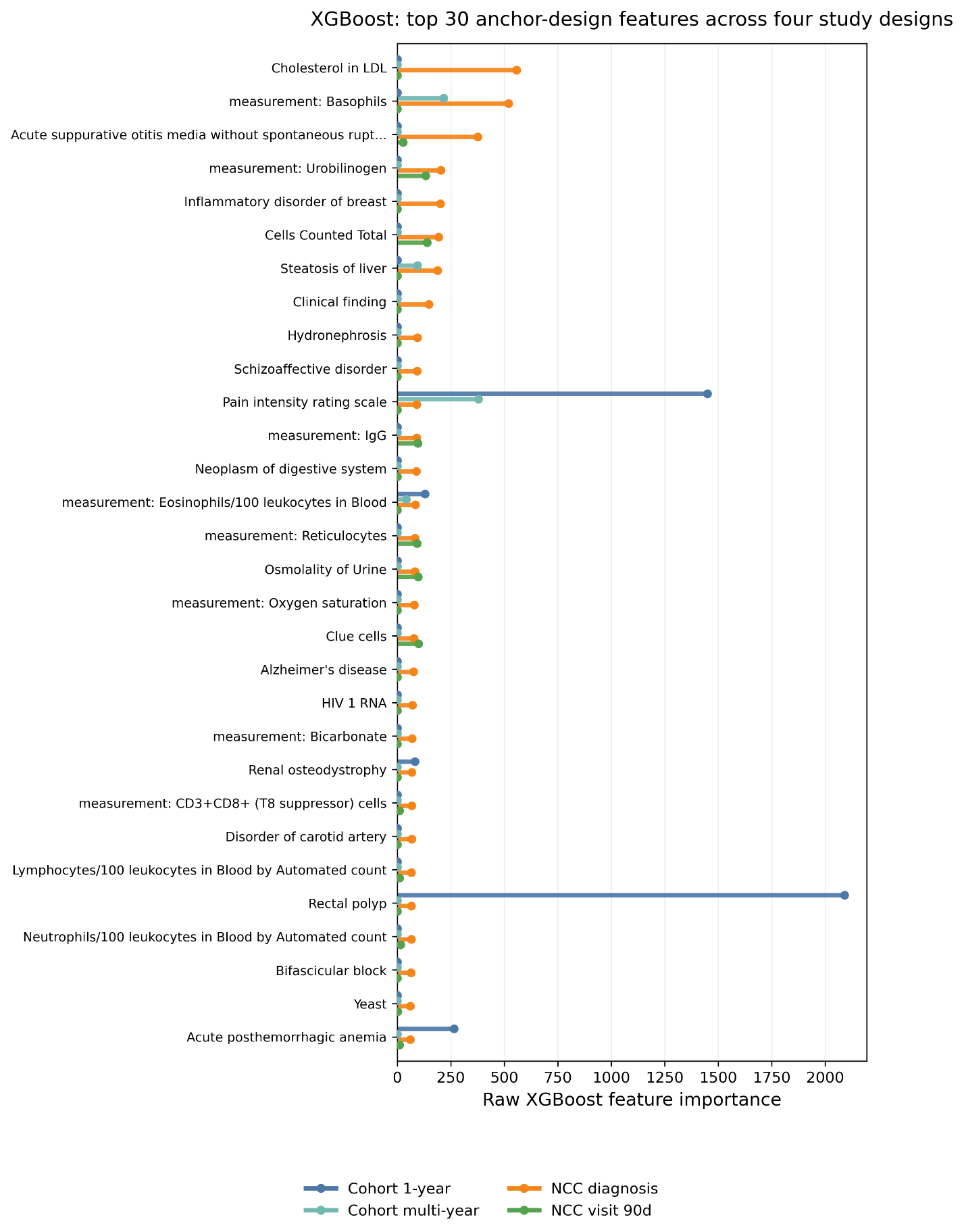

**Supplementary Figure 7. Top 30 MLP features ranked by attribution magnitude in Cohort-MultiYr.**

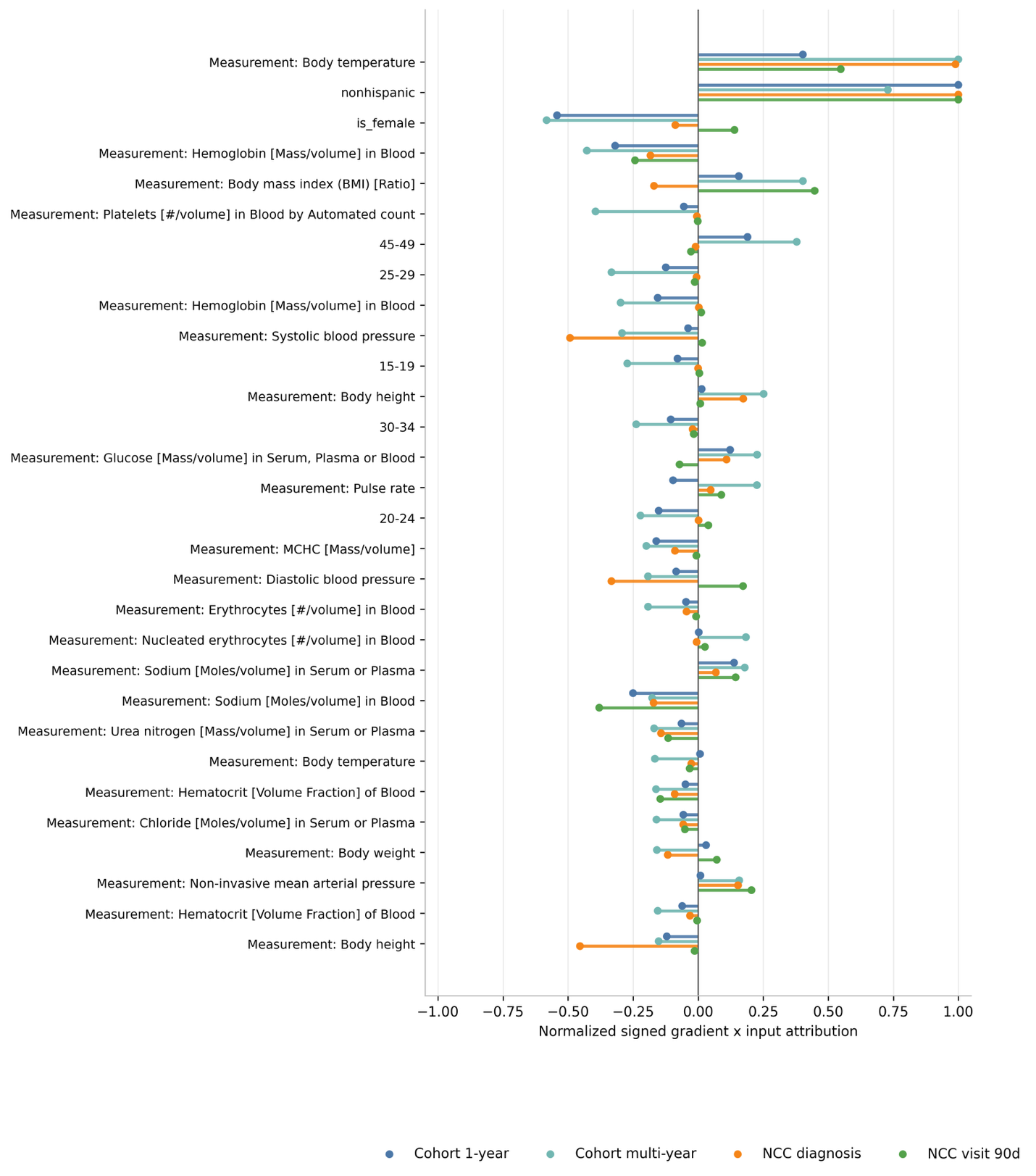
